# RNA sequencing of a large number of psoriatic patients identifies 131 novel miRNAs and 11 miRNAs associated with disease severity

**DOI:** 10.1101/2021.01.27.21250590

**Authors:** Å.Ø. Solvin, K. Chawla, L.C. Olsen, K. Danielsen, M. Jenssen, A.S. Furberg, M. Saunes, K. Hveem, P. Sætrom, M. Løset

## Abstract

**Background:** MicroRNAs are small regulatory molecules that are dysregulated in psoriasis. Despite previous efforts, much is unknown about the regulatory mechanisms of psoriasis genetics and their contributions to disease development and activity.

**Objectives:** To globally characterize the miRNAome of psoriatic skin in a large sample of psoriatic cases and controls for increased understanding of psoriasis pathophysiology.

**Methods:** Skin biopsies from psoriatic cases (n=75) and non-psoriatic controls (n=57) were RNA sequenced. Count data was meta-analyzed with a previously published dataset (cases, n=24, controls, n=20), increasing the number of psoriatic cases four-fold from previously published studies. Differential expression analyses were performed comparing lesional psoriatic (PP), non-lesional psoriatic (PN) and control (NN) skin. Further, functional enrichment and cell specific analyses were performed.

**Results:** We identified 439 significantly differentially expressed miRNAs (DEMs), of which 131 were novel and 11 were related to disease severity. Meta-analyses identified 20 DEMs between PN and NN, suggesting an inherent change in all psoriatic skin. By integrating the miRNA transcriptome with mRNA target interactions, we identified several functionally enriched terms, including ‘thyroid hormone signaling’, ‘insulin resistance’ and various infectious diseases. Cell specific expression analyses revealed that the upregulated DEMs were enriched in epithelial and immune cells.

**Conclusions:** We have provided the most comprehensive overview of the miRNome in psoriatic skin to date and identified a miRNA signature related to psoriasis severity. Our results may represent molecular links between psoriasis and related comorbidities and have outlined potential directions for future functional studies to identify biomarkers and treatment targets.

## Introduction

Psoriasis is a complex disease, triggered by environmental factors in genetically susceptible individuals. The prevalence is substantially higher in the Nordic countries ^1-3^ compared to global estimates ^4^. Genomic studies have established psoriasis as a Th-17/IL-23 dominated disease ^5^ and guided development of novel treatments ^5^. However, the exact pathophysiological mechanisms are unknown, and the regulatory mechanisms of psoriasis genetics have not yet been fully explained.

MicroRNAs (miRNAs) are non-coding RNAs, contributing to post-transcriptional downregulation of gene expression by repressing or degrading their target messenger RNAs (mRNAs) ^6^. More than 60% of human mRNAs contain evolutionary conserved miRNA target sites, suggesting that miRNAs can have a profound impact on cellular states and disease phenotypes ^7^. The first psoriasis-specific miRNAs were published in 2007 ^8^. With increasing sample sizes, advancing technologies and analytical methods, several differentially expressed miRNAs (DEMs) have since been identified ^8-14^. Targeted characterization has shown that many of these DEMs play central roles in regulating cellular functions and miRNAs have been suggested as sensitive biomarkers for disease activity, prognosis and treatment response ^15^. However, previous studies have involved a relatively small number of skin samples, some did not include a control group and the majority have employed microarrays rather than next-generation sequencing (NGS). For the study with the largest reported sample size to date (psoriatic cases, n = 24) ^11^, we found the reproducibility of the reported miRNAs unclear, most likely due to their choice of statistical approach.

To comprehensively profile the psoriasis miRNAome, we used high-resolution RNA sequencing (RNAseq) of a large number of lesional psoriatic (PP), non-lesional psoriatic (PN) and control skin (NN) skin from 121 individuals (cases, n = 75, controls, n = 46). To increase the power to detect novel DEMs, we did a robust statistical re-analysis of the largest dataset published to date (GSE31037)^11^ and meta-analyzed this with our data. The meta-analysis included 99 psoriatic cases and 66 non-psoriatic controls, increasing the number of psoriatic cases four-fold from previously published studies on miRNAs in psoriasis.

### Patients and methods

#### Participants and skin samples

Participants were adults of European descent. Psoriatic cases had psoriasis vulgaris and underwent a wash-out period without any topical or systemic-/photoherapy for two and four weeks prior to participation, respectively. Controls did not have psoriasis or any other inflammatory skin disease, nor any first-degree relatives with psoriasis. Disease severity was assessed using Psoriasis Area and Severity Index (PASI). From cases, two 4mm skin biopsies were collected; one PP and one PN. From controls one 4mm biopsy NN was collected. If possible, the PN and NN samples were taken from a sun-protected area on the buttock. Biopsies were snap frozen in liquid nitrogen and later stored at − 80 °C. Written informed consent was obtained from all participants. The study was approved by the Regional Committee for Medical and Health Research Ethics in Mid-Norway and conducted according to the Declaration of Helsinki principles.

### Total RNA extraction, library preparation and RNA sequencing

Skin biopsies were homogenized using Precellys homogenizer (Bertin Technologies, USA) and RNA was extracted using mirVana Isolation Kit (Applied Biosystems, Foster City, CA, USA). RNA integrity (RIN) was assessed using the Agilent RNA 6000 Nano kit on a 2100 Bioanalyzer chip (Agilent Technologies, Inc., Santa Clara, CA, USA) and the mean RIN was 8.2. Sequencing libraries were prepared using the NEXTflex small RNA-seq kit *v*3 (Bio Scientific, Austin, TX, USA). Single-end read sequencing was performed on an Illumina HiSeq4000 instrument, in accordance with the manufacturer’s instructions (Illumina, Inc., San Diego, CA, USA). The reads were mapped to human miRNAs in the miRBase database *v*22.1 ^16^ with the miraligner tool ^17^.

### Differential expression analysis

We identified DEMs in our samples (henceforth; the NTNU dataset) across three contrasts (PP versus NN, PP versus PN and PN versus NN) using a linear model adjusted for sex, age and BMI. The count matrices were transformed using the voom algorithm from the limma package of Bioconductor ^18,19^. The Benjamini-Hochberg method ^20^ was used to control the false discovery rate (FDR), and miRNAs with FDR <0.05 were considered significantly differentially expressed. We reanalyzed the hitherto largest published miRNA dataset in psoriatic skin (NCBI GEO accession number: GSE31037) (henceforth; the Joyce dataset)^11^, using the same statistical approach as on the NTNU dataset before meta-analysis. The meta-analysis included an additional factor estimating expression differences between the two datasets in the linear model. To determine if any miRNAs were related to psoriasis severity, cases were separated into mild (PASI <10) and moderate/severe (PASI ≥10) disease before differential expression analysis.

### Target prediction and functional enrichment analysis

Target genes for the top 20 most up- and downregulated DEMs were identified using the TargetScan database ^21^. Functional enrichment of Kyoto Encyclopedia of Genes and Genomes (KEGG) terms was performed using the gProfileR package in R ^22^. DEMs associated with psoriasis severity were analyzed for enrichment of Reactome terms ^23^ using the online tool miRnet (www.mirnet.ca) ^24,25^.

### Cell and tissue specific expression analysis

The FANTOM5 atlas ^26^ was used to determine which cells express the top 10 most up- and downregulated DEMs by fold change (FC) in the meta-analysis, both overall and after PASI stratification.

## Results

### Study participants

A total of 75 psoriatic cases and 46 controls were enrolled in the study. Clinical characteristics are presented in Table 1.

**Table 1.** Clinical characteristics of psoriatic cases and controls in the NTNU dataset.

|  | *Cases (n = 75)* | *Controls (n = 46)* |
| --- | --- | --- |
| **Age, years** (SD) | 52.7 (16.0) | 38.9 (15.4) |
| **Women, n (%)** | 33 (44.0) | 23 (50.0) |
| **Height, cm** (SD) | 174.2 (9.1) | 176.9 (8.9) |
| **Weight, kg** (SD) | 86.4 (16.2) | 78.3 (14.9) |
| **BMI, kg/m<sup>2</sup>** (SD) | 28.5 (5.2) | 24.9 (3.8) |
| **Waist circumference, cm** (SD) | 100.3 (15.8) | 89.4 (12.0) |
| **PASI, AU** (SD) | 6.2 (5.3) | - |

SD, standard deviation; cm, centimeter; kg, kilogram; BMI, body mass index; m, meter; PASI, psoriasis area and severity index.

### Differentially expressed miRNAs

In PP/NN, we identified 375 DEMs in the NTNU dataset, 231 DEMs in the Joyce dataset and 395 DEMs in the meta-analysis (FDR <0.05, log_2_FC >0) (Table 2, supplementary table (ST) 1-3). In PP/PN, we identified 415, 225 and 401 DEMs in the NTNU, Joyce and meta-analysis datasets, respectively. In PN/NN, we identified four DEMs in the NTNU dataset and none in the Joyce dataset. However, when data was pooled for the meta-analysis, 20 DEMs were identified (Figure 1, ST. 4). Table 3 a-c shows the top 10 DEMs for each contrast in the meta-analysis.

**Table 2.** Differentially expressed miRNAs (log_2_FC >0, FDR <0.05) between PP/NN, PP/PN and PN/NN in the NTNU dataset, the Joyce dataset and the meta-analyzed dataset.

| **<u>Contrast</u>** | **Number of differentially expressed miRNAs** |  |  |
| --- | --- | --- | --- |
|  | *NTNU* | *Joyce* | *Meta-analysis* |
| **PP/NN** | 375 | 231 | 395 |
| **PP/PN** | 415 | 225 | 401 |
| **PN/NN** | 4 | 0 | 20 |

PP, lesional psoriatic skin; PN, non-lesional psoriatic skin; NN, healthy control skin

**Table 3.** Top 10 differentially expressed miRNA by FC in the meta-analysis for (**a**) PP/NN, (**b**) PP/PN and (**c**) PN/NN.

| miRNA | Log<sub>2</sub>FC | *P*-value | FDR |
| --- | --- | --- | --- |
| hsa-miR-31-3p | 4.238 | 7.210 x10<sup>-31</sup> | 1.668 x10<sup>-29</sup> |
| hsa-miR-31-5p | 3.948 | 8.900 x10<sup>-51</sup> | 9.469 x10<sup>-49</sup> |
| hsa-miR-21-3p | 2.697 | 3.708 x10<sup>-82</sup> | 1.973 x10<sup>-79</sup> |
| hsa-miR-6510-3p | -2.227 | 4.538 x10<sup>-46</sup> | 3.018 x10<sup>-44</sup> |
| hsa-miR-1268a | 2.171 | 1.992 x10<sup>-26</sup> | 2.864 x10<sup>-25</sup> |
| hsa-miR-1268b | 2.157 | 5.653 x10<sup>-26</sup> | 7.646 x10<sup>-25</sup> |
| hsa-miR-885-5p | -2.144 | 5.247 x10<sup>-51</sup> | 6.979 x10<sup>-49</sup> |
| hsa-miR-940 | 2.129 | 6.437 x10<sup>-30</sup> | 1.317 x10<sup>-28</sup> |
| hsa-miR-3617-5p | -2.020 | 1.380 x10<sup>-25</sup> | 1.669 x10<sup>-24</sup> |
| hsa-miR-33b-3p | 1.972 | 1.983 x10<sup>-23</sup> | 1.918 x10<sup>-22</sup> |

b)
| miRNA | Log<sub>2</sub>FC | *P*-value | FDR |
| --- | --- | --- | --- |
| hsa-miR-31-5p | 3.799 | 1.471 x10<sup>-57</sup> | 1.957 x10<sup>-55</sup> |
| hsa-miR-31-3p | 3.719 | 7.753 x10<sup>-33</sup> | 1.528 x10<sup>-31</sup> |
| hsa-miR-21-3p | 2.603 | 6.084x 10<sup>-91</sup> | 3.237 x10<sup>-88</sup> |
| hsa-miR-3617-5p | -2.068 | 2.225 x10<sup>-30</sup> | 3.199 x10<sup>-29</sup> |
| hsa-miR-135b-5p | 1.949 | 7.824 x10<sup>-59</sup> | 1.387 x10<sup>-56</sup> |
| hsa-miR-33b-3p | 1.944 | 1.858 x10<sup>-30</sup> | 2.746 x10<sup>-29</sup> |
| hsa-miR-940 | 1.913 | 4.166 x10<sup>-34</sup> | 9.234 x10<sup>-33</sup> |
| hsa-miR-4517 | 1.906 | 5.583 x10<sup>-38</sup> | 1.650 x10<sup>-36</sup> |
| hsa-miR-664b-3p | -1.899 | 2.954 x10<sup>-54</sup> | 2.619 x10<sup>-52</sup> |
| hsa-miR-885-5p | -1.853 | 1.662 x10<sup>-46</sup> | 1.263 x10<sup>-44</sup> |

c)
| miRNA | Log<sub>2</sub>FC | *P*-value | FDR |
| --- | --- | --- | --- |
| hsa-miR-6510-3p | -0.499 | 5.340 x10<sup>-5</sup> | 0.004 |
| hsa-miR-598-3p | -0.428 | 2.352 x10<sup>-5</sup> | 0.003 |
| hsa-miR-450a-5p | -0.411 | 1.708 x10<sup>-6</sup> | 4.54 x10<sup>-4</sup> |
| hsa-miR-214-3p | 0.373 | 2.747 x10<sup>-5</sup> | 0.003 |
| hsa-miR-34a-5p | 0.355 | 1.199 x10<sup>-7</sup> | 6.379 x10<sup>-5</sup> |
| hsa-miR-141-3p | -0.343 | 1.517 x10<sup>-5</sup> | 0.003 |
| hsa-miR-429 | -0.336 | 1.589 x10<sup>-4</sup> | 0.009 |
| hsa-miR-574-5p | 0.334 | 0.001 | 0.028 |
| hsa-miR-542-3p | -0.304 | 1.639 x10<sup>-4</sup> | 0.009 |
| hsa-miR-485-3p | 0.302 | 3.027 x10<sup>-4</sup> | 0.015 |

PP, lesional psoriatic skin; PN, non-lesional psoriatic skin; NN, healthy control skin; FC, fold change.

**Figure 1.**
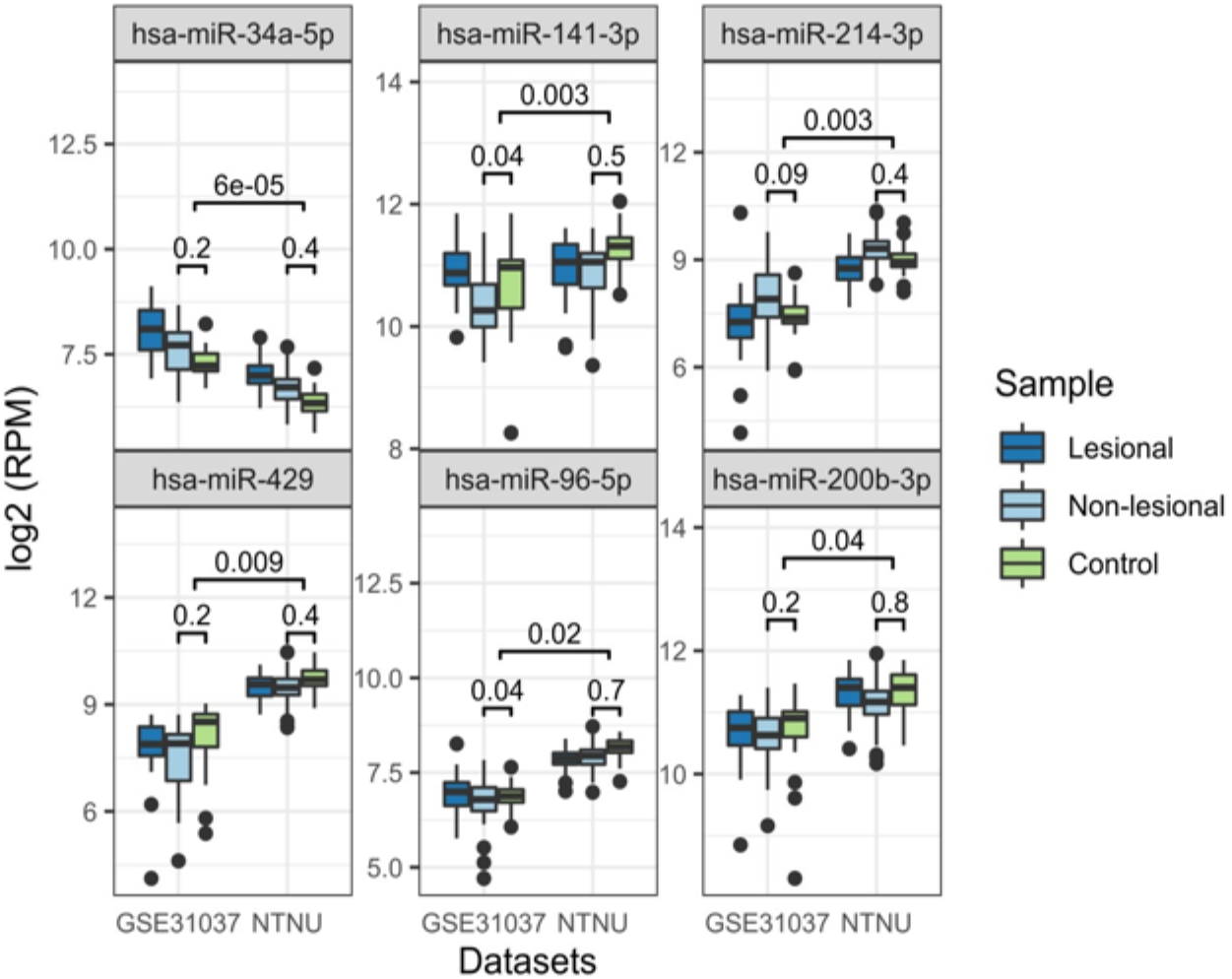
Box plot of expression levels of the six DEMs in PN/NN (meta-analysis) with reads per million (RPM) >100 in the NTNU and Joyce (GSE31037) data sets with *P*-values given for both separate and meta-analysis. These DEMs had small, but consistent expression levels across the datasets.

### Correlation between datasets

Shared DEMs in the two datasets were highly correlated (Pearson’s correlation coefficient r = 0.89 for PP/NN, r = 0.87 for PP/PN). Intersecting the DEMs showed substantial overlap (Figure 2a) and shared DEMs were mostly consistently expressed (Figure 2b). A total of 28 DEMs in PP/NN were exclusively identified in the meta-analysis and many of these were among the DEMs with the lowest log_2_FC (Supplementary figure 1, ST. 5)

**Figure 2.**
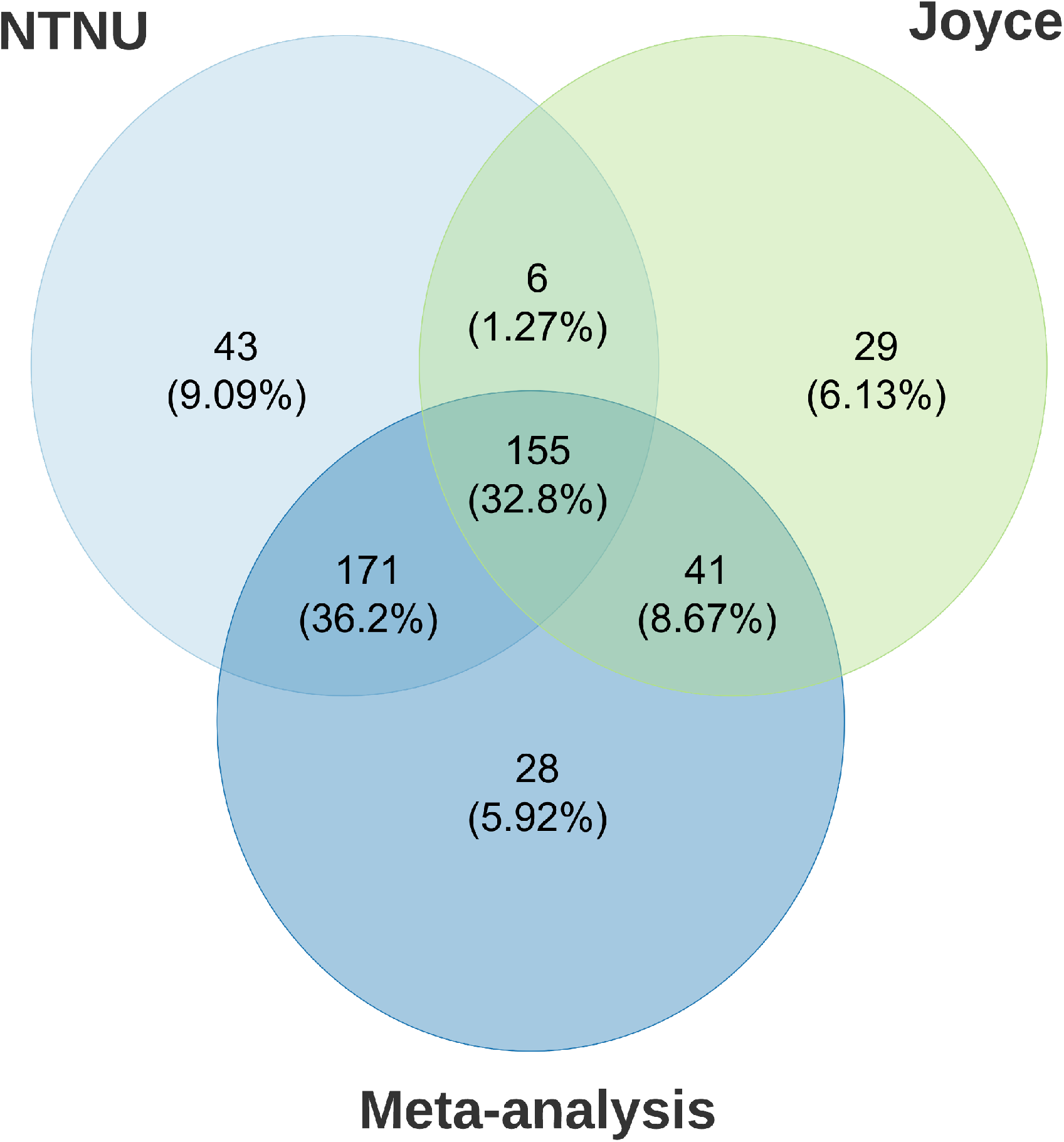
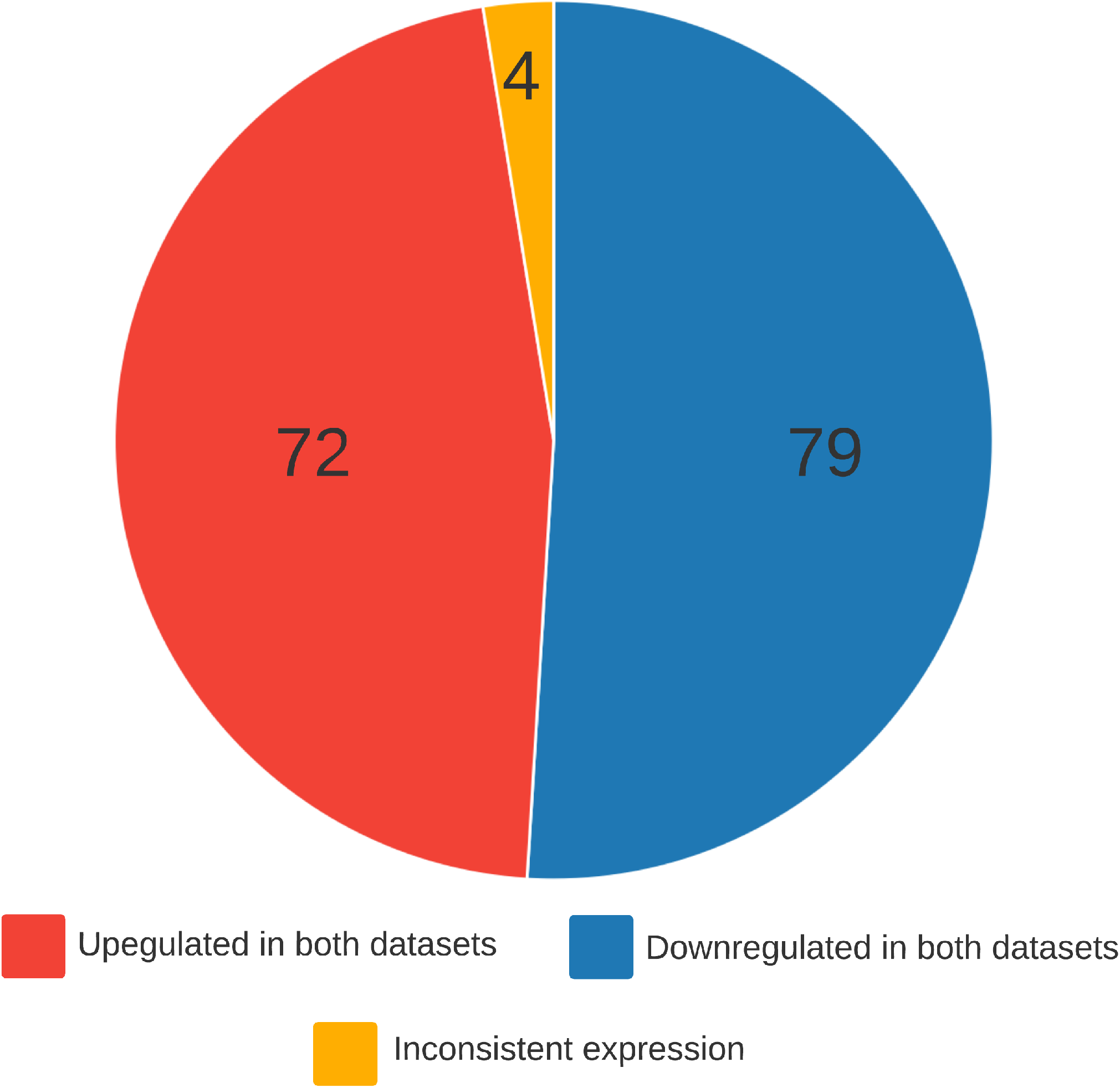
**a)** Venn-diagram showing overlap of the DEMs between datasets for PP/NN at FDR <0.05. A total of 155 (33%) of DEMs were identified in all three datasets. **b)** Pie chart of the direction of fold change of the shared DEMs between all three datasets in PP/NN. Of the 155 DEMs, 151 (97%) exhibit consistent expression patterns in the Joyce and NTNU datasets, and only four DEMs were inconsistently expressed.

### Novel miRNAs

To identify novel miRNAs in psoriatic skin we compared our results from the meta-analysis to previously published studies ^9-12,14^. In total, we identified 131 novel miRNAs (ST. 6), corresponding to 33% of the DEMs in the meta-analysis.

### Biological functions

To deduce the possible biological functions of the DEMs, targets of the miRNAs with average expression ≥100 reads per million (RPM) and FDR <0.05 for PP/NN and PN/NN in the meta-analyzed data set were identified using TargetScan ^21^. This threshold was chosen as most miRNAs below this threshold show no functional activity ^27,28^. We are not presenting results from PP/PN, as these were similar to PP/NN. Predicted targets in PP/NN were enriched for several terms (Figure 3), including ‘Thyroid hormone signaling pathway’, ‘insulin resistance’, ‘human papillomavirus infection’ and ‘melanogenesis’. The most significantly enriched term for PP/NN and PN/NN was ‘MicroRNAs in cancer’.

**Figure 3.**
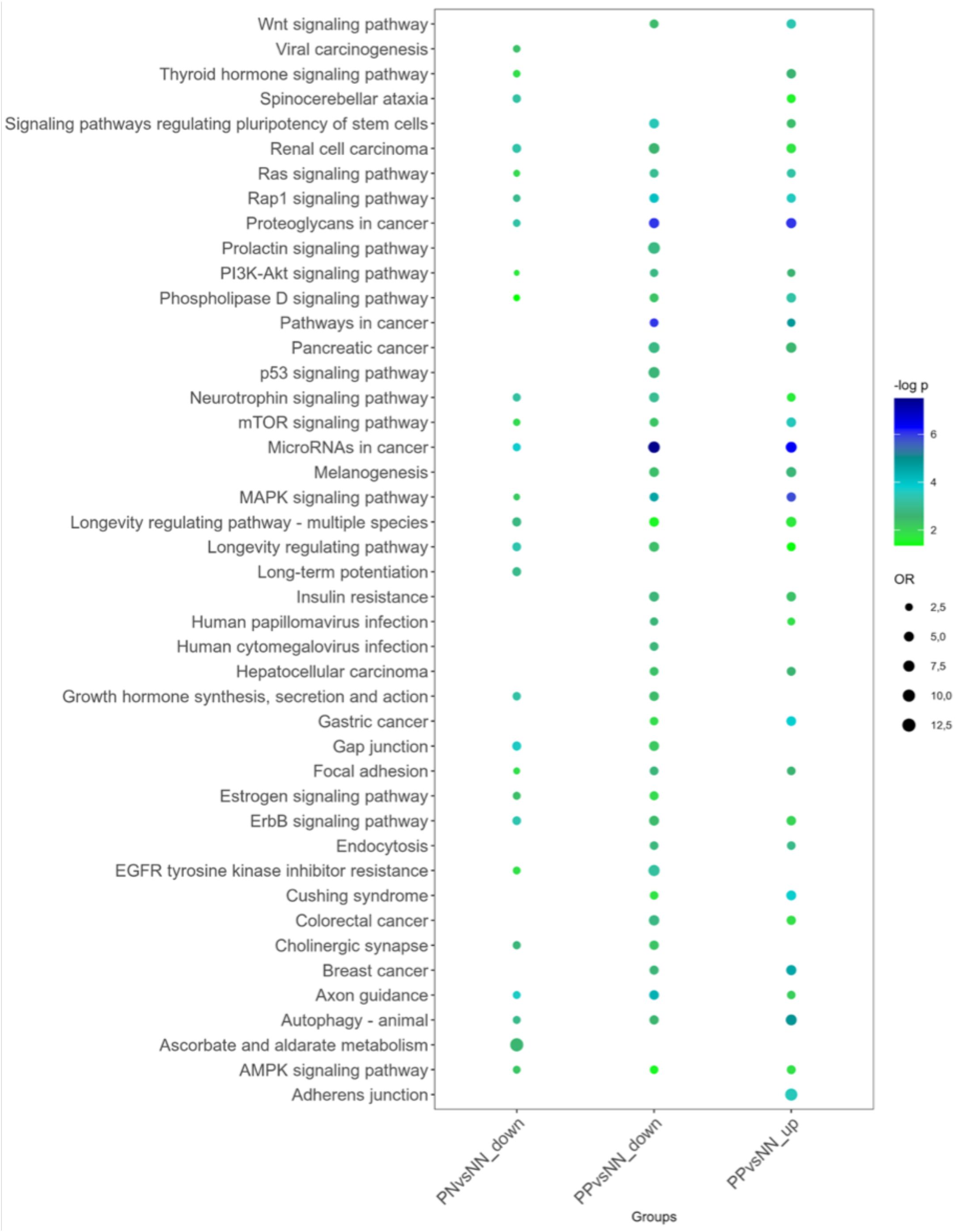
Dot plot of the most significantly enriched KEGG pathways among the targets of up-and downregulated DEMs for PP/NN and PN/NN in the meta-analysis. Abbreviations: Wnt, wingless-related integration site; Rap1, Ras-related protein 1; PI3K-Akt, phosphoinositide-3 kinase-akt; mTOR, mechanistic target of rapamycin; MAPK, mitogen-activated protein kinase; EGFR, epidermal growth factor; AMPK, AMP-activated protein kinase.

### Cell and tissue specific expression

To evaluate which cells that express the top 10 up- and downregulated DEMs by FC in PP/NN and PN/NN, we extracted cell specific data from the FANTOM5 atlas ^26^. In PP/NN DEMs were enriched in fibroblasts and epithelial cells, as well as in immune cells including monocytes and leukocytes (Figure 4). In PN/NN, several of the DEMs were enriched in epithelial and endo-epithelial cells (Figure 4).

**Figure 4.**
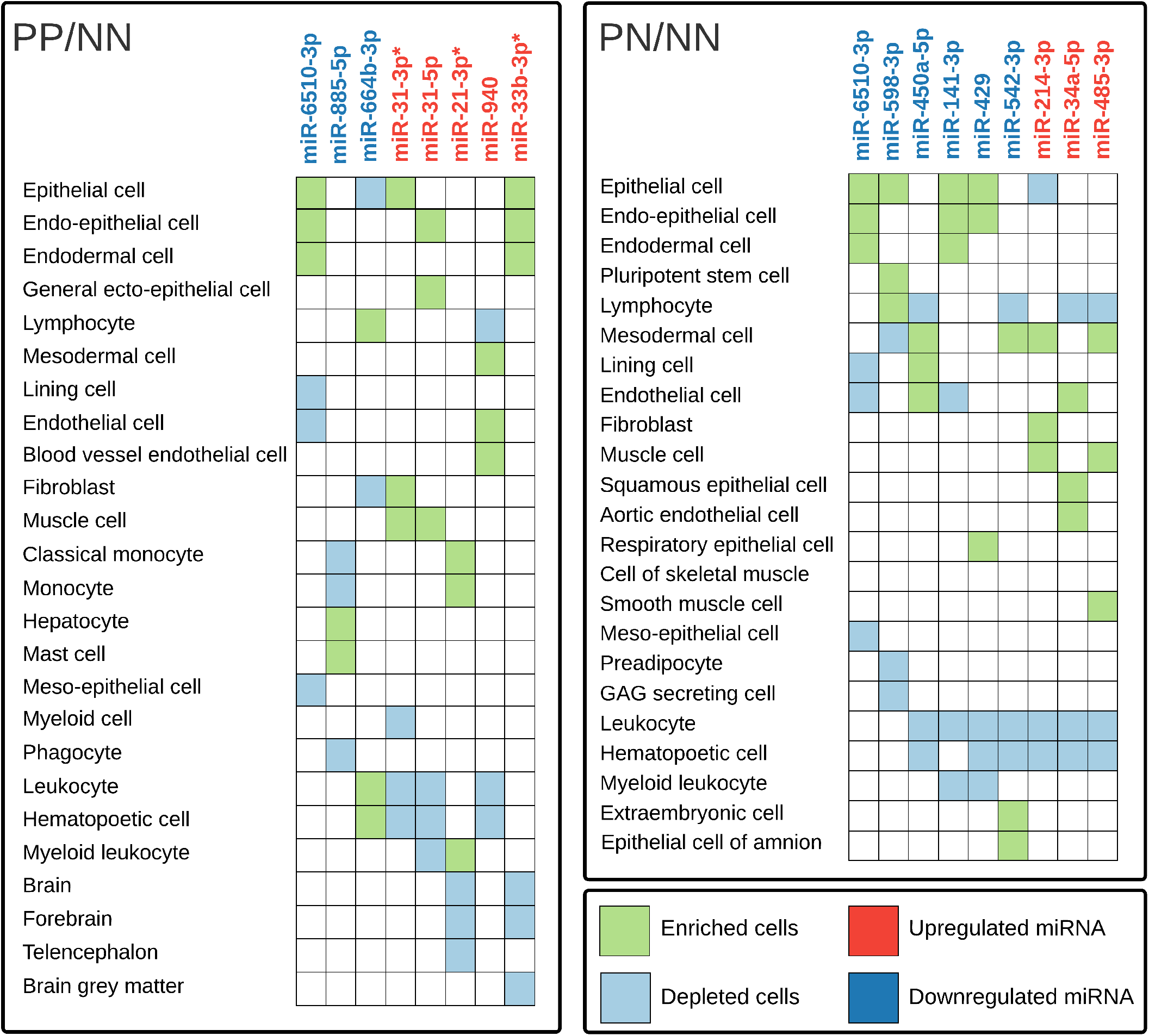
Primary cell expression analysis using the FANTOM5 atlas for the top 10 differentially expressed miRNAs (DEMs) in the meta-analysis in PP/NN and PN/NN. The miRNAs with an asterisk∗ were analyzed based on pre-miRNA, as the mature miRNA was not annotated. Abbreviations: miRNA, micro RNA; GAG, glycosaminoglycan.

Cell specific KEGG enrichment analysis showed that the enriched terms for each cell type were largely overlapping with the enriched terms for non-cell specific DEMs (Figures 3, 5). There was no clear separation of enriched terms between the different cell types.

**Figure 5.**
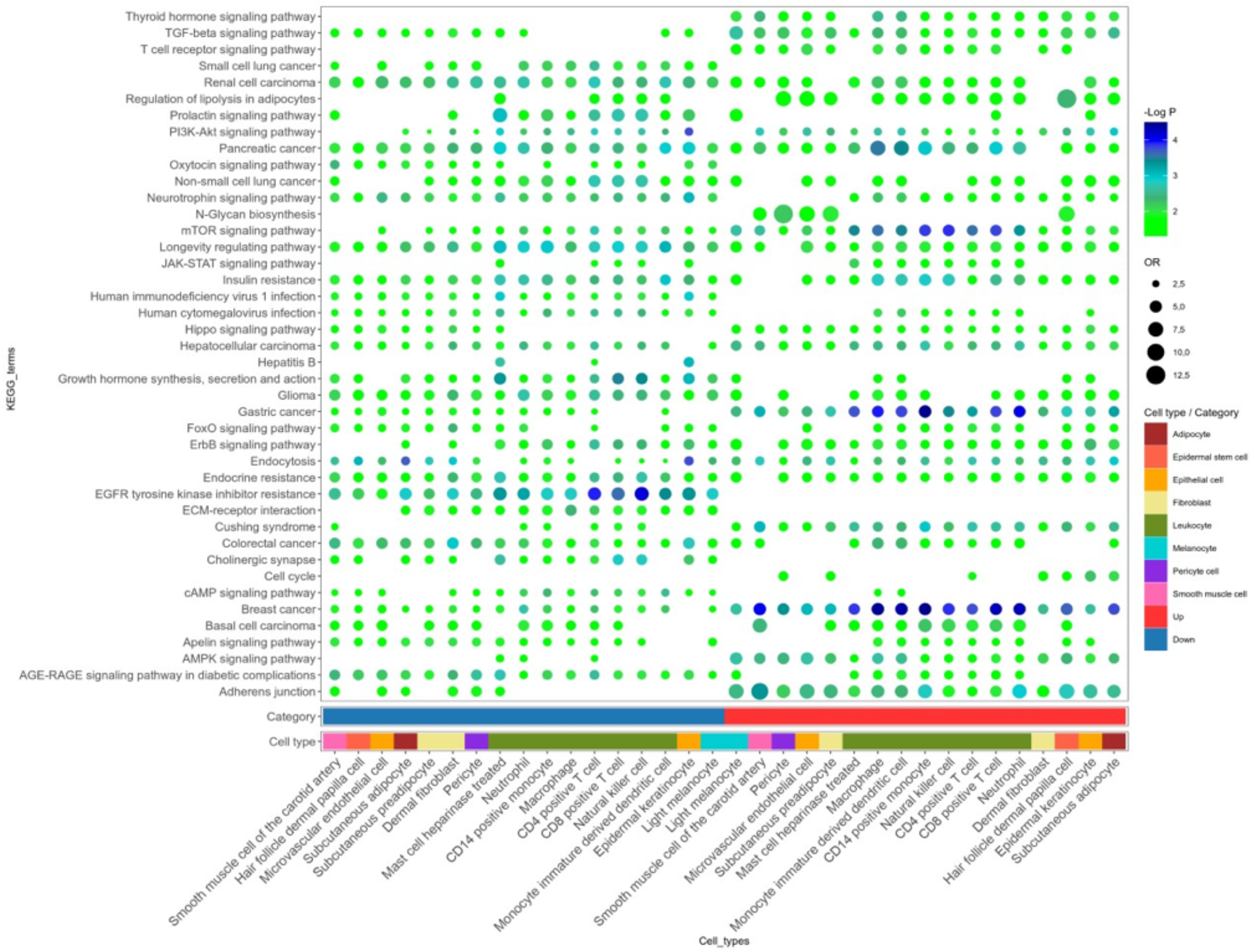
Enrichment analysis of DEMs by cell type showed a large overlap of enriched terms for up- and downregulated DEMs. Colors on the bottom bar indicate cell type and color on second bottom bar indicate direction of DEM regulation. Certain terms were only enriched for up-, and downregulated DEMs, including ‘epidermal growth factor receptor (EGFR)’, ‘tyrosine kinase inhibitor resistance’, ‘thyroid hormone signaling pathway’ and ‘T cell receptor signaling pathway’. ‘Hepatitis B’ was only enriched for upregulated DEMs in leukocytes. Terms enriched in leukocytes were more significant as a group than the other cell types. Abbreviations: TGF, transforming growth fatcor; PI3K-Akt, phosphoinositide-3 kinase-akt; mTOR, mechanistic target of rapamycin; JAK-STAT, janus kinase-signal transducer and activator of transcription; FoxO, forkhead box transcription factor class O; EGFR, epidermal growth factor; ECM, extracellular matrix; cAMP, cyclic adenosine monophosphate; AMPK, AMP-activated protein kinase; AGE-RAGE, advanced glycation end products-receptor for advanced glycation end products; CD, cluster of differentiation; OR, odds ratio.

### Psoriasis severity specific miRNAs

We dichotomized the cases into mild (PASI <10) and moderate/severe (PASI ≥10) and identified five DEMs in PP skin and six DEMs in PN skin. In PP skin, all five miRNAs were upregulated, while in PN skin, four miRNAs were upregulated and two were downregulated (ST. 7). The targets of the upregulated DEMs in PP and PN skin were enriched for inflammatory pathways including ‘IL1-signaling’, ‘transforming growth factor beta (TGFβ)-receptor complex’ and toll-like receptor (TLR) cascades. The targets of the downregulated DEMs were enriched in mRNA processing pathways including splicing and translation. miR-374a and miR-146a were both upregulated in PP skin and these were enriched in myeloid leukocytes and hematopoietic cells (Figure 6). All DEMs in PN were depleted in leukocytes and hematopoietic cells.

**Figure 6.**
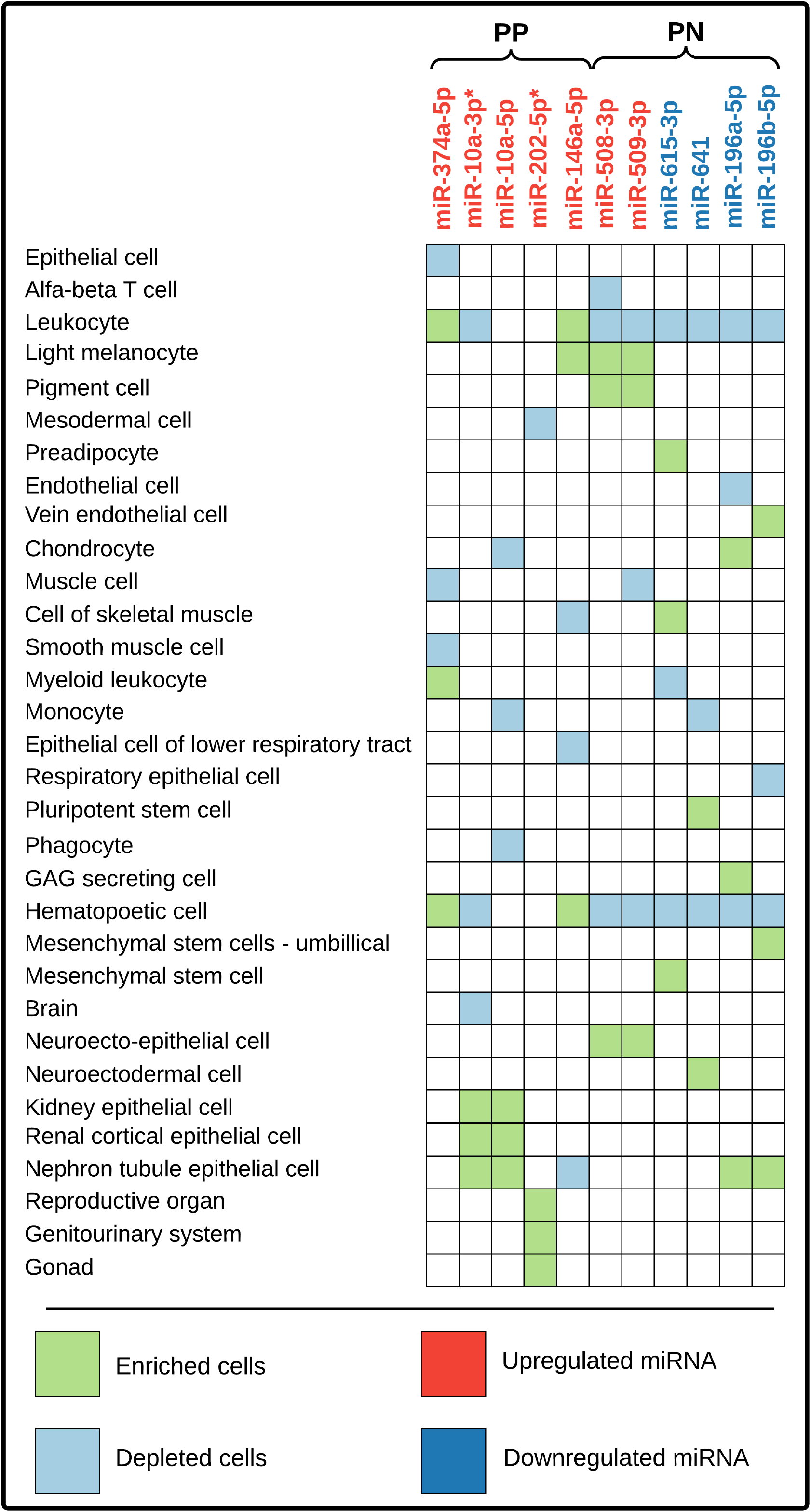
Primary cell expression analysis using the FANTOM5-atlas for the DEMs identified in moderate/severe versus mild psoriasis. The miRNAs with an asterisk∗ were analyzed based on pre-miRNA, as the mature miRNA was not annotated. Abbreviations: GAG, glycosaminoglycan; miRNA, micro RNA.

## Discussion

In this study, we explored the miRNAome in psoriatic skin by analyzing RNAseq data from 99 psoriatic cases and 66 controls, increasing the number of psoriatic cases four-fold from previous work. By improving statistical power due to a large sample, we identified 439 DEMs, of which 131 were novel and 11 were related to psoriasis severity. Further, we identified 20 DEMs between psoriatic non-lesional (PN) and healthy control skin (NN). This suggests an inherent change in all psoriatic skin, supporting the previously shown ‘pre-inflammatory’ signature outside established psoriatic plaques for protein-coding RNAs ^29^. We identified enrichment of terms representing potential disease pathology (e.g. ‘Wnt-’ and ‘Ras signaling pathway’) and established or emerging comorbidities (e.g. ‘insulin resistance’ and ‘thyroid hormone signaling’).

We replicated several of the most known miRNAs in psoriatic skin, including miR-21, miR-31 and miR-146a ^30^. In PP/NN, miR-31-5p and mir-31-3p were the most differentially expressed by FC. miR-31 has been shown to increase keratinocyte proliferation and migration and targets epithelial membrane protein 1 (EMP-1) ^31^ and E2F transcription factor 2 (E2F2), a known cell cycle regulator promoting proliferation ^32^. miRNA-21, overexpressed in PP/NN, regulates keratinocyte proliferation and differentiation ^15^ as well as T-cell apoptosis ^33^. miR-128a was also highly upregulated in PP/NN. This miRNA is a regulator in Wnt-, and IL-23 signaling and is upregulated in peripheral blood neutrophils of patients with diffuse cutaneous systemic sclerosis ^34^. This miRNA also targets genes involved in insulin signaling and may mediate some of the increased risk of insulin resistance and obesity in psoriasis ^35^. While the majority of the DEMs were upregulated, some were downregulated, including miR-6510-3p (log_2_FC −2.23). This miRNA is also downregulated in oral lichen planus ^36,37^, an inflammatory and proliferative disease of the oral mucosa, as well as in squamous cell carcinoma ^37^. Taken together, we have replicated known miRNAs in psoriasis and expanded the psoriatic miRNAome by identifying 131 novel DEMs.

The DEMs in PN/NN skin had lower FCs compared to PP/NN, however, they reflect inherent changes in all psoriatic skin and could play important roles in disease pathology. The upregulated miRNAs miR-34a-5p and miR-214-3p, target common genes (Figure 7) involved in cell cycle regulation and apoptosis. miR-34a has also been shown to promote keratinocyte inflammation ^38^. Interestingly, three of the downregulated DEMs in PN/NN (miR-429, −200b and −141) are part of the miR-200 family. This miRNA family is highly expressed in skin and is central for skin homeostatic functions ^39^. These miRNAs also have shared target genes (*MALT1* and *ZEB1*) involved in NF-kB activation and signaling, as well as in T-cell receptor signaling. Our PN/NN results support the finding of an inflammatory and proliferative response in skin outside of established plaques ^29^.

**Figure 7.**
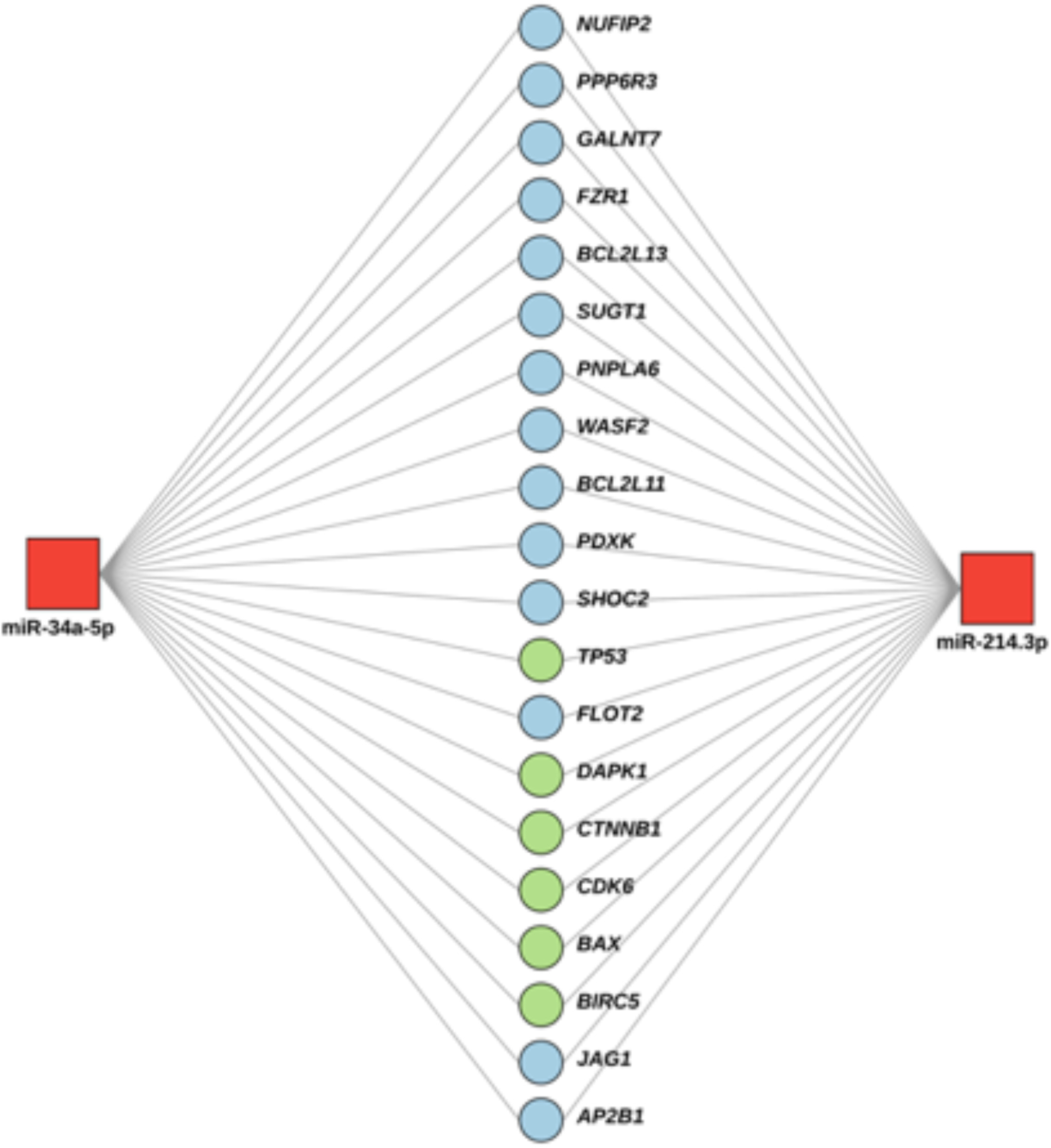
Upregulated miRNAs in PN/NN and their shared targets. Genes in green are part of the KEGG term ‘pathways in cancer’. Target interactions and the illustration were created using miRnet^24,25^.

Enrichment analysis identified terms involved in well-known disturbed functions in psoriasis pathology. This included functions crucial for epidermal- and stem cell behavior and development (‘Wnt signaling’) ^40^, epidermal proliferation and differentiation (‘Ras signaling’) ^41^ and viral infections (e.g. ‘human papillomavirus infection’). The term ‘Hepatitis B’ was significantly enriched for downregulated DEMs in PP/NN. This term contains genes important for TNF- and NF-kB signaling and leukocyte migration. Examples include CAMP responsive element binding protein 1 (*CREB1*), mitogen-activated protein kinase 8 (*MAPK8)*, B-cell lymphoma 2 (*BCL2*) and phosphatidylinositol-4,5-bisphosphate 3-kinase catalytic subunit alpha (*PIK3CA*), which are targets of 38, 26, 22 and 25 of the DEMs, respectively. This proposes these DEMs as mediators of inflammation regulation in psoriasis. Both within the melanogenesis and viral infection terms, there were also genes important for specific inflammatory processes associated with psoriasis, e.g. tumor necrosis factor (TNF) ^42^, Th17-cell differentiation, and IL-17 signaling. Some of the genes are targets of many DEMs, including *TGFBR1* and *MAPK1*, targeted by 26 and 24 DEMs, respectively. Various cancer terms were also enriched for the downregulated miRNAs. These terms contain genes important for cell cycle and vascular endothelial growth factor (VEGF) signaling, e.g. *PIK3CA*, AKT serine/threonine kinase (*AKT*) 3 and cyclin dependent kinase (*CDK*) 6. These genes are targets of 25, 26 and 28 downregulated DEMs, respectively, suggesting that the miRNAs exert their functions through these target interactions. Some enriched pathways may also represent a shared molecular basis for psoriasis comorbidities, such as thyroid hormone signaling and insulin resistance.

‘MicroRNAs in cancer’ was the most significantly enriched term for both PP/NN and PN/NN. This term contains cell cycle regulators such as E2F3 and CDK6, potentially reflecting increased proliferation in both PP and PN skin. In PN/NN, the targets were enriched for pathways important for proliferation and inflammation, including ‘MAPK-signaling’ and ‘ErbB-signaling’, demonstrating inherent changes in all skin of psoriatic patients. Further, the downregulated miRNAs in PN/NN and PP/NN were enriched for ‘axon guidance’. Axonal function has been recognized as an important factor in pruritus ^43^, a symptom of psoriasis often overlooked.

In this study, we obtained full-thickness skin biopsies, containing a mixture of cell types, including keratinocytes, immune cells, fibroblasts and skin appendages. By extracting data from the FANTOM5 atlas ^26^, we aimed to dissect the cell specific origin of the DEMs. The top DEMs in PP/NN were enriched in cell types important for skin structure, such as epithelial cells and fibroblasts and immune cells such as monocytes and leukocytes. We further aimed to elucidate the functions of the DEMs by cell type. Almost all cell types expressed miRNAs whose targets were enriched for insulin resistance, supporting the hypothesis of systemic inflammation and shared inflammatory pathways as the tie between psoriasis and metabolic syndrome/cardiovascular disease ^35^. We acknowledge the limitations of using the FANTOM5 atlas for cell specific analysis as it is based on a limited set of non-inflamed skin samples. Further, our cell specific analyses focused on highly expressed miRNAs, which may downplay effects of miRNAs in rare cell types. To circumvent the dilution of low-abundant, but important transcripts, single-cell RNAseq (scRNA-seq) can be applied. scRNA-seq of a small number of psoriatic samples have identified the important role of endothelial cells expressing leukocyte adhesion molecules^44^ as well as trajectories for keratinocyte differentiation ^45^.

Patients with severe psoriasis show a higher risk of comorbidities ^46^ and the underlying molecular mechanisms may differ according to severity. We identified 11 DEMs comparing moderate/severe to mild psoriasis. The upregulated DEMs in both PP and PN skin were enriched for inflammatory pathways. The downregulated DEMs were enriched for pathways involved in mRNA processing, such as splicing and translation. This suggests that the upregulated DEMs reflect or respond to changes in immune function, while the downregulated DEMs are involved in more basic cellular functions. To determine if these changes are a cause or a consequence of increasing disease severity, functional studies are needed. Cell specific expression of the severity associated DEMs identified miRNAs enriched in both leukocytes and hematopoietic cells, suggesting that these migrate to the site upon plaque formation. Hematopoietic cells have been implicated in regulation of proliferating skin tissues during an early inflammatory phase ^47^.

A major strength of this study is the large number of cases and controls. Our study provides insight into the effect of applying different statistical methods on gene expression data. The Joyce data (GSE31037) was originally analyzed using a Pearson’s chi-squared test of goodness of fit on the normalized total reads from all PP, PN and NN skin ^11^. To the best of our knowledge, this approach is not beneficial to identify significant expression differences between sample groups, as it ignores individual variation within each sample group and thereby inflates the significance of highly expressed genes with higher variation than lowly expressed genes ^19^. We therefore reanalyzed the Joyce dataset which reduced the number of DEMs from the originally reported 317 to 257 (FDR<0.05, FC >1.4).

miRNA target prediction methods (including TargetScan), base predictions on statistically enriched sequence patterns in 3’ UTRs for genes that are down- and upregulated in experiments where individual miRNAs are respectively up- and downregulated. Consequently, these methods identify hypothetical interactions between miRNAs and mRNAs. However, the relationship between miRNA expression and mRNA repression is not linear and depends on a variety of factors including sequence complementarity and alternative polyadenylation ^21,48^. As such, the in vivo effects of endogenous miRNAs are difficult to predict reliably.

In conclusion, this study provides comprehensive insights into the miRNAome of psoriatic skin. By including a large sample of psoriatic cases and controls, we have identified 131 novel miRNAs for psoriasis, 11 miRNAs for psoriasis severity, and validated previously known psoriasis-associated miRNAs. We identified 20 DEMs in PN/NN, representing inherent alterations in the miRNAome of all skin of psoriatic patients. This advocates for the systemic involvement of the disease and may represent shared molecular mechanisms of the systemic inflammation of psoriasis. Our study outlines future directions for targeted functional follow-up studies to explore the causal effects of the DEMs. Increased knowledge of the psoriasis transcriptome can prove crucial in understanding disease pathology, which can catalyze breakthroughs in the development of biomarkers for diagnostic purposes, predicting comorbidities and treatment response, as well as identifying treatment targets.

## Data Availability

Due to Norwegian privacy laws, sequencing data will not be uploaded to a publicly available repository. Count-data will be made available upon request.

## Acknowledgements

We are grateful to all the volunteers who contributed to our study as well as to assistance from staff at the Clinical Research Facility, St. Olavs hospital, Trondheim University Hospital, Norway and Biobank1, Trondheim, Norway. The total RNA extraction, library preparation and RNAseq were performed in close collaboration with the Genomics Core Facility (GCF), at the Norwegian University of Science and Technology (NTNU). The GCF is funded by the F. aculty of Medicine and Health Sciences at NTNU and Central Norway Regional Health Authority. This work was supported by the Liaison Committee between the Central Norway Regional Health Authority and NTNU.

## Notes

**Conflicts of interest** None declared.

**Funding sources** ML, PS, KH, KC and LCO work in a research unit funded by Stiftelsen Kristian Gerhard Jebsen; Faculty of Medicine and Health Sciences, NTNU; The Liaison Committee for education, research and innovation in Central Norway; and the Joint Research Committee between St. Olavs hospital and the Faculty of Medicine and Health Sciences, NTNU. ML was supported by a research grant from the Liaison Committee for education, research and innovation in Central Norway. The funders had no influence on study design, data collection and analysis, decision to publish, or preparation of the manuscript.

### Competing Interest Statement

The authors have declared no competing interest.

### Funding Statement

ML, PS, KH, KC and LCO work in a research unit funded by Stiftelsen Kristian Gerhard Jebsen; Faculty of Medicine and Health Sciences, NTNU; The Liaison Committee for education, research and innovation in Central Norway; and the Joint Research Committee between St. Olavs hospital and the Faculty of Medicine and Health Sciences, NTNU. ML was supported by a research grant from the Liaison Committee for education, research and innovation in Central Norway. The funders had no influence on study design, data collection and analysis, decision to publish, or preparation of the manuscript.

### Author Declarations

Regional Etichal Committee of Mid-Norway

