## Supplementary figure for "RNA sequencing of a large number of psoriatic patients identifies 131 novel miRNAs and 11 miRNAs associated with disease severity"

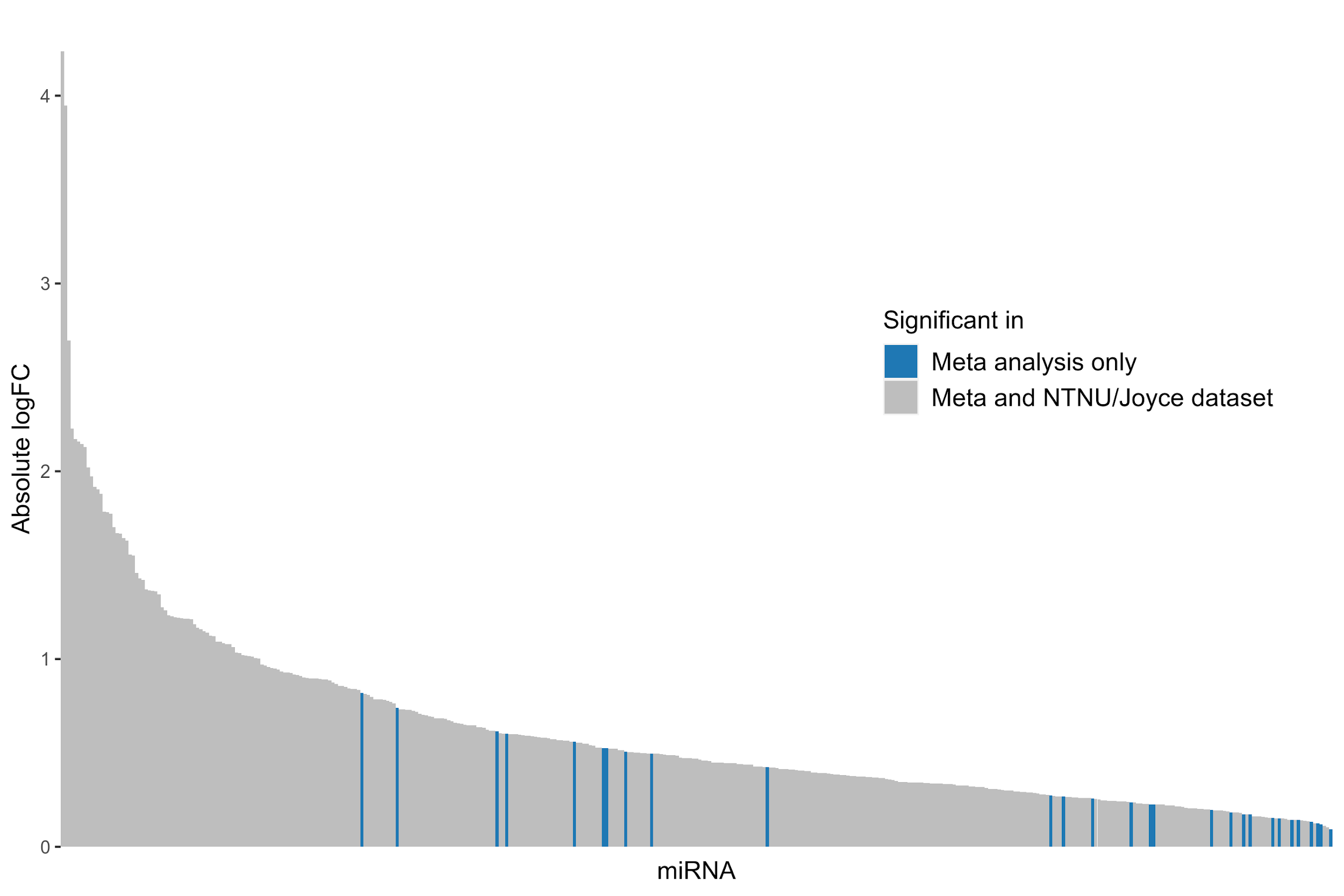

Supplementary figure 1: Absolute expression of significant miRNAs in PP/NN from the meta-analysis. The miRNAs which were only identified in the meta-analysis are highlighted in blue. The remaining miRNAs in grey are identified in at least one of the other datasets.
